# Lung cancer in the French West Indies: role of sugarcane work and other occupational exposures

**DOI:** 10.1101/2022.07.20.22277823

**Authors:** Léïla Cabréra, Aviane Auguste, Léah Michineau, Clarisse Joachim, Jacqueline Deloumeaux, Danièle Luce

## Abstract

**Objective:** To study the role of occupational exposures in lung cancer risk in the French West Indies, with special attention to some specific activities, such as sugarcane work, that can only be studied in a limited number of populations.

**Methods:** We used data from a population-based case-control study that included 147 incident lung cancer cases and 405 controls. Smoking history and a detailed occupational history with a description of tasks and substances were collected by questionnaire during face-to-face interviews. Odds ratios (OR) adjusted for sex, age, region, smoking status and cigarette pack-years and 95% confidence intervals (95% CI) were estimated by unconditional logistic regression.

**Results:** Significantly increased risks of lung cancer were found in sugarcane farm workers (OR=2.7; 95% CI 1.1-6.6) and more generally in the sugarcane growing sector (OR=2.5; 95% CI 1.0-6.3) and to a lesser extent in rum production. Elevated risks of lung cancer were also observed among other agricultural workers, painters, warehouse porters, labourers, maintenance and motor vehicle repair workers. Exposure to herbicides in sugarcane cultivation was associated with an increased risk of lung cancer (OR=2.6; 95% CI 0.9-7.6).

**Conclusion:** These results showed that occupational exposures contributed to lung cancer risk in the French West Indies, and highlighted the role of exposures related to sugarcane work.

## INTRODUCTION

Lung cancer is the leading cancer in the world in terms of incidence and mortality. In 2018, it is estimated that there were nearly 2.1 million new cases of lung cancer and 1.8 million deaths[1].

The main risk factor for lung cancer is smoking, which is responsible for at least 8 out of 10 lung cancer cases in Western countries. The risk of lung cancer development is around 10 times higher in smokers than in non-smokers, and the risk increases with the duration of smoking and the amount of cigarettes smoked, for all histological types of lung cancer[2].

While smoking is the major risk factor, many other risk factors for lung cancer have been identified. A large proportion of these are substances or exposure situations present mainly in the workplace, and lung cancer is the most common occupational cancer. Several occupational exposures are known risk factors for lung cancer, including asbestos, crystalline silica, diesel exhaust, polycyclic aromatic hydrocarbons (PAHs), various metals (arsenic, cadmium, beryllium, some chromium and nickel compounds), welding fumes, and ionising radiation. There is remaining uncertainty with regard to several probable occupational lung carcinogens, such as bitumen or non-arsenical insecticides[3].

Guadeloupe and Martinique, known as the French West Indies (FWI), are French overseas territories in the Caribbean with a population of around 800,000, mostly Afro-Caribbean. The age-standardised (world) lung cancer incidence rates per 100,000 over the period 2007-2016 are 12.1 in Guadeloupe and 10.3 in Martinique for men; for women these rates are respectively 4.4 and 6.3[4].

This relatively low incidence is mainly due to low tobacco consumption[5]. Indeed, the proportion of current smokers in the FWI is 23% among men and 13% among women. The proportion of those who have ever smoked tobacco is 38% among men and 19% among women. This population with a low prevalence of smoking is therefore of particular interest for the study of other risk factors for lung cancer, foremost among which are occupational exposures. The involvement of risk factors other than tobacco is also suggested by a descriptive study on lung cancers in Guadeloupe[6], which showed a high proportion of non-smoking cases.

The FWI population presents particularities in terms of occupational risks, with specific activities such as banana cultivation and the sugarcane industry. It has been suggested that there is excess lung cancer among banana plantation workers[7]. Elevated risks of lung cancer have also been found in sugarcane cultivation or the sugarcane industry[8]. However, the number of studies is limited and these results need to be replicated in other appropriate populations. As in other tropical regions, the FWI are also characterised by a significant use of pesticides. Apart from these specific exposures, the high proportion of very small enterprises (more than 90%) and the high frequency of informal employment are associated with poor working conditions.

The role of occupational exposures in the aetiology of lung cancer in the Caribbean remains largely unknown. Therefore, the objective was to examine the associations between occupational exposures and lung cancer risk in the FWI and to clarify the impact of certain tasks and substances on lung cancer risk.

## METHODS

This work is based on data from a population-based case-control study conducted in Guadeloupe and Martinique between 2013 and 2016. This study represents an extension to the FWI of the ICARE study, a large case-control study on respiratory cancers conducted in mainland France[9], and used the same protocol and questionnaire, with some adaptations to the local context. The study included both lung cancer and head and neck cancer cases, with a common control group. The present analysis included only lung cancer cases and controls.

The cases were identified in collaboration with the cancer registries of the two regions. Eligible cases were all patients with a newly diagnosed primary tumour of the trachea, bronchus, and lung (International Classification of Diseases, 10th revision, codes C33-C34) diagnosed between April 1, 2013, and December 31, 2016, residing in Guadeloupe or Martinique, and aged 75 years or younger at diagnosis. All histological types were included. The control group was a random sample of the general population recruited by random digit dialling, including cell phones. Recruitment was stratified to obtain a distribution of controls by age, sex, and region comparable to that of all cases. Additional stratification was used to obtain a distribution by socio-professional category comparable to that of the population (obtained from census data), to control for potential selection bias resulting from differential participation rates by socio-professional category.

Of the 237 cases identified as potentially eligible for the study, 169 (71.3%) agreed to participate and were interviewed. Of these, after review of the diagnosis, 22 (13.0%) did not meet the inclusion criteria. Of the 497 eligible controls, 405 (81.5%) completed the questionnaire. A total of 147 incident lung cancer cases and 405 controls were included in the study, for a total of 552 subjects. Each subject included in the study gave written informed consent. Cases and controls were interviewed by specially trained interviewers, with a standardised questionnaire including socio-demographic characteristics, smoking habits, detailed occupational history, with a description of each job held (work position, company activity, tasks performed), and specific questionnaires for certain occupations or activities. These job-specific questionnaires included a comprehensive list of questions on exposures, tasks performed and materials handled. In addition to the job-specific questionnaires originally used in the ICARE study[9], we developed a questionnaire for the sugarcane industry and we modified the agriculture questionnaire to better describe local agricultural activities.

For each subject, all jobs performed during his or her working life (occupation and industry) were coded, using the International Standard Classification of Occupations (ISCO) of the International Labour Office of 1968 for the occupation, and the Nomenclature d’Activités Française (NAF) of the INSEE of 2000 for the industry. The coding was performed by a trained coder blind to case-control status. We then generated dichotomous variables for each occupation/industry: “having ever worked in a given occupation/industry” versus “having never worked in this occupation/industry” using three ISCO levels (1, 2 and 5-digit codes) and two NAF levels (2 and 4-digit codes). We were also interested in tasks and substances in the workplace known or suspected to be associated with an increased risk of lung cancer. Algorithms were created from the responses to the different questionnaires in order to assess exposure to asbestos, diesel or gasoline exhaust, fumes, dusts, solvents, or welding. The agriculture and sugarcane industry questionnaires were analysed in detail, to evaluate exposure to pesticides and several tasks related to sugarcane work. All exposures were evaluated in two categories: ever exposed vs. never exposed.

Unconditional logistic regression models were used to estimate adjusted odds ratios (ORs) and their 95% confidence intervals (95% CIs), and to assess interactions. ORs were adjusted for sex, age (continuous), region (Guadeloupe or Martinique), smoking status (never smoker, ex-smoker, current smoker), and cumulative cigarette quantity in pack-years (continuous). Never smokers are those who have smoked less than 100 cigarettes in their lifetime. Ex-smokers are those who quit smoking at least two years previously.

## RESULTS

In our study, as expected, the proportion of ever smokers was higher in the cases than in the controls (60% vs. 34% respectively) and the risk of lung cancer was significantly increased in ex-smokers (OR=2.5; 95% CI 1.4-4.3) and in current smokers (OR=8.5; 95% CI 4.8-14.8). Examination of the distribution of cases according to histological type showed a high proportion of adenocarcinomas (62%), followed by squamous cell carcinomas (19%) and other histological types (19%).

Table 1 presents the data related to the study of occupations performed by the subjects. For the three ISCO code levels, ORs are presented only for occupations with at least five exposed cases. Overall, the ORs for professional, technical and related workers, clerical and related workers and sale workers (ISCO codes 0/1 to 4) were less than 1. However, a slight increase in risk was observed for service workers (ISCO code 5), agricultural, animal husbandry and forestry workers, fishermen and hunters (ISCO code 6) and production and related workers, transport equipment operators and labourers (ISCO code 7/8/9).

**Table 1.**
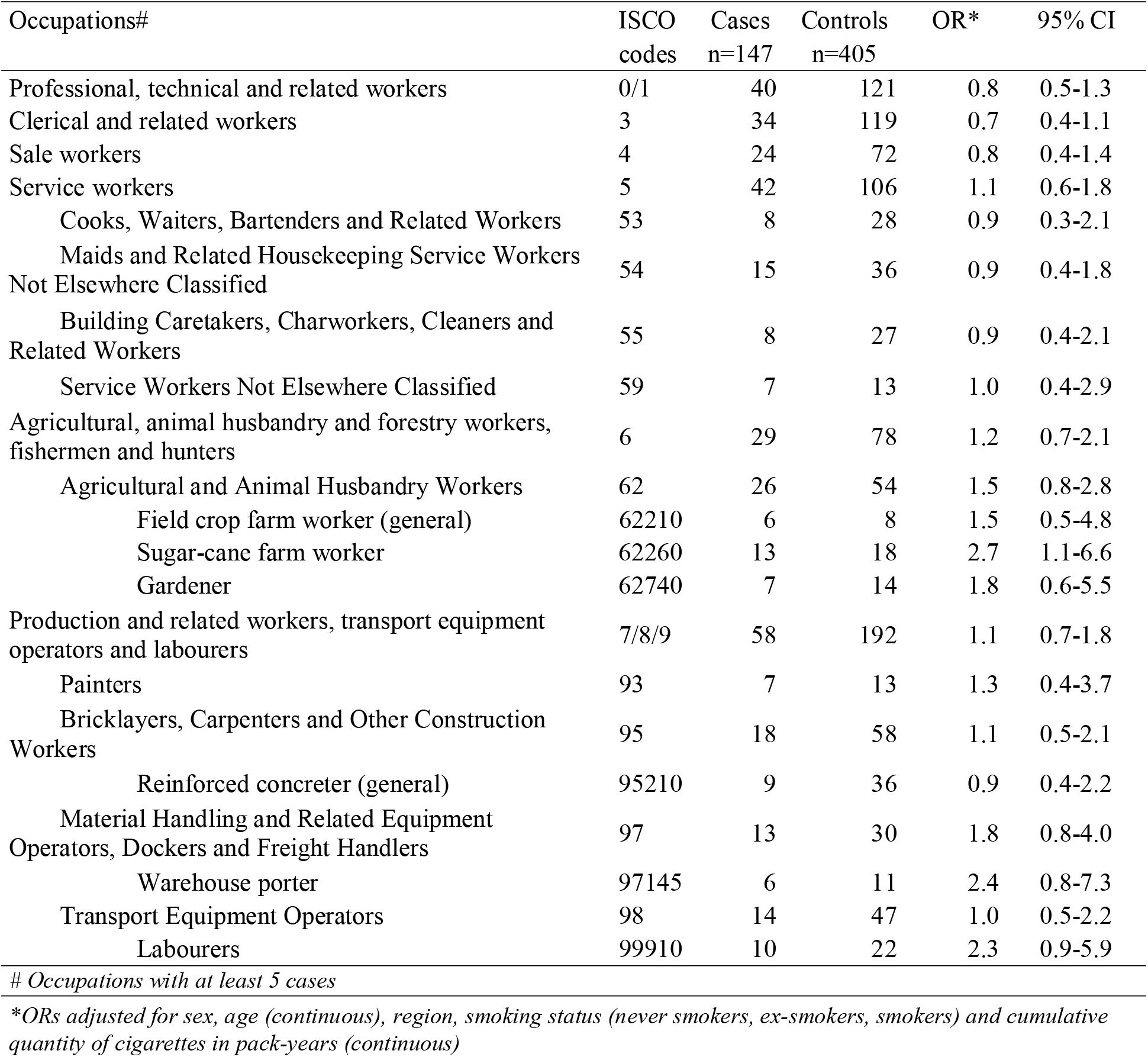
Association between selected occupations and lung cancer risk in the French West Indies (FWI)

Looking in more detail at the next ISCO level, 2-digit codes, borderline significant ORs greater than 1 were observed for agricultural and animal husbandry workers (ISCO code 62) and labourers not elsewhere classified (ISCO code 99), 1.5 and 2.3 respectively. High, although not significant, ORs were also observed for painters (ISCO code 93), bricklayers, carpenters and other construction workers (ISCO code 95) and material handling and related equipment operators, dockers and freight handlers (ISCO code 97).

A more detailed analysis for agricultural and animal husbandry workers showed a significantly increased risk (OR=2.7; 95% CI 1.1-6.6) for sugarcane farm workers (ISCO code 62260), and a moderate increase in risk for gardeners (ISCO code 62740) and field crop farm workers (ISCO code 62210). It should be noted that this code includes banana workers, who cannot be clearly identified by ISCO. The increase in risk observed for the category of “material handling and related equipment operators, dockers and freight handlers” only concerned warehouse porters (ISCO code 97145). The labourers subgroup included only one occupation. For bricklayers, carpenters and other construction workers, no high OR was observed at the 5-digit ISCO level with at least five exposed cases.

ORs associated with the different industries (2 and 4-character NAF codes) are presented in Table 2, for industries with at least five exposed cases. A non-significant increase in lung cancer risk was observed in agriculture, hunting and related service activities (NAF code 01). This increase was limited to the growing of cereals and other crops n.e.c. and the growing of fruit. The growing of cereals and other crops n.e.c (NAF code 011A) corresponded here exclusively to sugarcane cultivation, for which a significant OR of 2.5 was observed (OR=2.5; 95% CI 1.0-6.3), consistent with the high OR found for sugarcane farm workers. The growing of fruit (NAF code 011F) corresponded to banana cultivation, associated with a non-significant OR of 1.1. The OR of 1.7 associated with the manufacture of distilled potable alcoholic beverages (NAF code 159A, here, rum manufacturing) was consistent with the risk increases observed for other sugarcane workers. High but non-significant ORs were also observed in the maintenance and repair of motor vehicles (NAF code 502Z), the wholesale and retail trade (NAF codes 51 and 52, respectively), general (overall) public service activities (NAF code 751A), primary education (NAF code 801Z) as well as technical and vocational secondary education (NAF code 802C), and finally other service activities (NAF code 93) and activities of households as employers of domestic staff (NAF code 950Z).

**Table 2.**
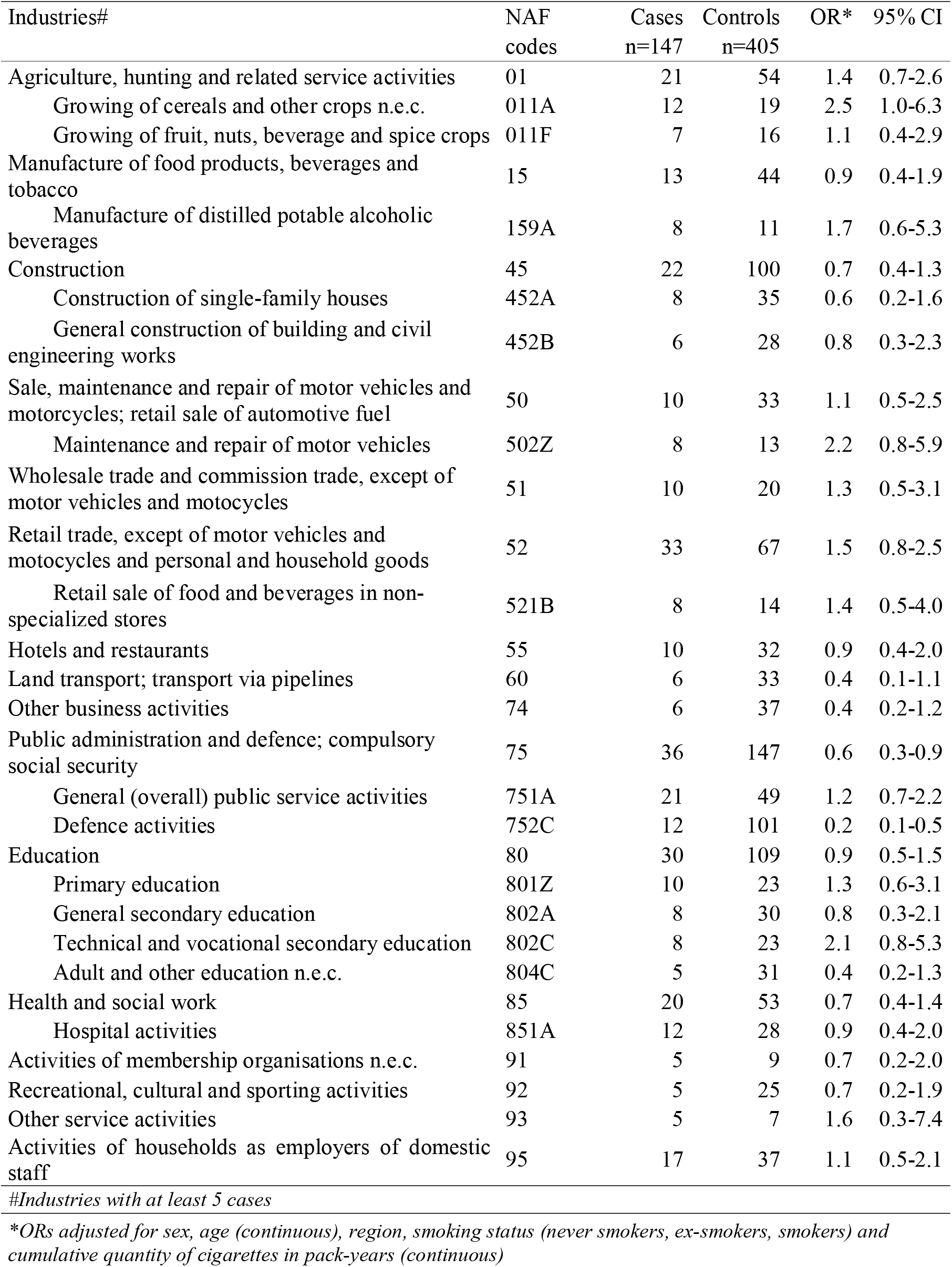
Association between selected industries and lung cancer risk in the French West Indies

| Industries# | NAF codes | Cases<br>n=147 | Controls<br>n=405 | OR* | 95% CI |
| --- | --- | --- | --- | --- | --- |
| Agriculture, hunting and related service activities | 01 | 21 | 54 | 1.4 | 0.7-2.6 |
| Growing of cereals and other crops n.e.c. | 011A | 12 | 19 | 2.5 | 1.0-6.3 |
| Growing of fruit, nuts, beverage and spice crops | 011F | 7 | 16 | 1.1 | 0.4-2.9 |
| Manufacture of food products, beverages and tobacco | 15 | 13 | 44 | 0.9 | 0.4-1.9 |
| Manufacture of distilled potable alcoholic beverages | 159A | 8 | 11 | 1.7 | 0.6-5.3 |
| Construction | 45 | 22 | 100 | 0.7 | 0.4-1.3 |
| Construction of single-family houses | 452A | 8 | 35 | 0.6 | 0.2-1.6 |
| General construction of building and civil engineering works | 452B | 6 | 28 | 0.8 | 0.3-2.3 |
| Sale, maintenance and repair of motor vehicles and motorcycles; retail sale of automotive fuel | 50 | 10 | 33 | 1.1 | 0.5-2.5 |
| Maintenance and repair of motor vehicles | 502Z | 8 | 13 | 2.2 | 0.8-5.9 |
| Wholesale trade and commission trade, except of motor vehicles and motorcycles | 51 | 10 | 20 | 1.3 | 0.5-3.1 |
| Retail trade, except of motor vehicles and motorcycles and personal and household goods | 52 | 33 | 67 | 1.5 | 0.8-2.5 |
| Retail sale of food and beverages in non-specialized stores | 521B | 8 | 14 | 1.4 | 0.5-4.0 |
| Hotels and restaurants | 55 | 10 | 32 | 0.9 | 0.4-2.0 |
| Land transport; transport via pipelines | 60 | 6 | 33 | 0.4 | 0.1-1.1 |
| Other business activities | 74 | 6 | 37 | 0.4 | 0.2-1.2 |
| Public administration and defence; compulsory social security | 75 | 36 | 147 | 0.6 | 0.3-0.9 |
| General (overall) public service activities | 751A | 21 | 49 | 1.2 | 0.7-2.2 |
| Defence activities | 752C | 12 | 101 | 0.2 | 0.1-0.5 |
| Education | 80 | 30 | 109 | 0.9 | 0.5-1.5 |
| Primary education | 801Z | 10 | 23 | 1.3 | 0.6-3.1 |
| General secondary education | 802A | 8 | 30 | 0.8 | 0.3-2.1 |
| Technical and vocational secondary education | 802C | 8 | 23 | 2.1 | 0.8-5.3 |
| Adult and other education n.e.c. | 804C | 5 | 31 | 0.4 | 0.2-1.3 |
| Health and social work | 85 | 20 | 53 | 0.7 | 0.4-1.4 |
| Hospital activities | 851A | 12 | 28 | 0.9 | 0.4-2.0 |
| Activities of membership organisations n.e.c. | 91 | 5 | 9 | 0.7 | 0.2-2.0 |
| Recreational, cultural and sporting activities | 92 | 5 | 25 | 0.7 | 0.2-1.9 |
| Other service activities | 93 | 5 | 7 | 1.6 | 0.3-7.4 |
| Activities of households as employers of domestic staff | 95 | 17 | 37 | 1.1 | 0.5-2.1 |

#Industries with at least 5 cases
\*ORs adjusted for sex, age (continuous), region, smoking status (never smokers, ex-smokers, smokers) and cumulative quantity of cigarettes in pack-years (continuous)

In addition to this analysis of job titles, a number of exposure situations known or suspected to be associated with an increased risk of lung cancer were further analysed (Table 3). Although not significant, high ORs were found for exposures to diesel and gasoline exhaust, paint, fumes, solvents, wood treatment products, and disinfection of agricultural premises. Of note, the OR for exposure to fumes was borderline significant (OR=1.4; 95% CI 0.9-2.3). Conversely, no association was found for exposure to dusts, acids, welding or asbestos.

**Table 3.** Association between occupational exposures and specific tasks and lung cancer risk in the French West Indies

| Exposures | Cases<br>n=147 | Controls<br>n=405 | OR* | 95% CI |
| --- | --- | --- | --- | --- |
| Diesel exhaust | 22 | 54 | 1.4 | 0.7-2.6 |
| Gasoline exhaust | 18 | 48 | 1.2 | 0.6-2.4 |
| Dusts | 66 | 235 | 0.8 | 0.5-1.2 |
| Fumes | 43 | 99 | 1.4 | 0.9-2.3 |
| Acids | 25 | 87 | 0.8 | 0.4-1.4 |
| Welding | 9 | 49 | 0.7 | 0.3-1.6 |
| Painting | 21 | 54 | 1.3 | 0.7-2.4 |
| Asbestos | 13 | 75 | 0.6 | 0.3-1.2 |
| Solvents | 24 | 60 | 1.3 | 0.7-2.4 |
| Wood treatment products | 9 | 15 | 1.5 | 0.5-4.1 |
| Other chemical products | 4 | 5 | 1.4 | 0.2-7.5 |
| Disinfection of agricultural premises | 4 | 11 | 1.6 | 0.4-7.0 |
| Pesticides in general<sup>a</sup> | 13 | 36 | 1.6 | 0.7-3.5 |
| Pesticides in sugar cane | 8 | 13 | 2.6 | 0.9-7.6 |
| In banana | 6 | 8 | 3.0 | 0.8-10.5 |
| In other crops | 4 | 21 | 1.2 | 0.4-3.8 |
| Insecticides in general | 4 | 17 | 1.2 | 0.4-4.0 |
| Insecticides in sugar cane | 0 | 0 | - | - |
| In banana | 1 | 6 | 0.7 | 0.1-6.9 |
| In other crops | 3 | 10 | 1.7 | 0.4-7.1 |
| Herbicides in general | 13 | 32 | 1.9 | 0.8-4.1 |
| Herbicides in sugar cane | 8 | 13 | 2.6 | 0.9-7.6 |
| In banana | 6 | 7 | 3.2 | 0.9-11.6 |
| In other crops | 4 | 16 | 1.7 | 0.5-5.6 |
| Fungicides in general | 2 | 18 | 0.8 | 0.2-3.6 |
| Fungicides in sugar cane | 0 | 0 | - | - |
| Fungicides in banana | 1 | 5 | 1.8 | 0.2-16.7 |
| Fungicides in other crops | 1 | 12 | 0.6 | 0.1-4.8 |
| Sugarcane work |  |  |  |  |
| Cutting of the sugarcane | 9 | 18 | 1.4 | 0.5-3.8 |
| Collecting the sugarcane | 6 | 16 | 1.8 | 0.6-5.4 |
| Burning of the sugarcane | 4 | 15 | 0.6 | 0.1-2.1 |
| Other work in the field | 4 | 14 | 0.9 | 0.2-3.6 |
| Crushing of the sugarcane | 2 | 1 | 3.7 | 0.1-125.3 |
| Handling the sugarcane | 10 | 20 | 1.9 | 0.7-4.7 |

\*ORs adjusted for sex, age (continuous), region, smoking status (never smokers, ex-smokers, smokers) and cumulative quantity of cigarettes in pack-years (continuous)
<sup>a</sup>Pesticides in general includes insecticides, herbicides and fungicides, all crops combined

We analysed occupational exposure to pesticides (i.e. herbicides, insecticides, fungicides and other biocides) in sugarcane cultivation, banana cultivation and in other crops. A strong, borderline significant increase in risk was associated with sugarcane pesticides (OR=2.6; 95% CI 0.9-7.6). This increase was entirely due to herbicides, as none of the subjects in our study reported applying either insecticides or fungicides to sugarcane. An increased risk (OR=3.0; 95% CI 0.8-10.5) was also found for banana pesticides. Again, this increase was mainly due to the application of herbicides (OR=3.2; 95% CI 0.9-11.6). Insecticides and herbicides used on other crops were associated with lower and non-significant ORs. No association was found with the use of fungicides.

We examined certain tasks related to sugarcane cultivation in greater detail (Table 3). The activities of cutting, collecting, and crushing sugarcane, as well as handling, were associated with a non-significant increase in lung cancer risk, with respective ORs of 1.4, 1.8, 3.7, and 1.9. No association was found with the burning of sugarcane before or after harvesting

## DISCUSSION

To our knowledge, this is the first study on the occupational risk factors for lung cancer in the Caribbean. As classically observed in lung cancer studies, the proportion of smokers was higher in cases than in controls, however in our study the proportion of cases who had never smoked amounted to 40% and was higher than the values usually observed, which are typically in the order of 10 to 20%. This means that in these regions other factors are indeed involved in the occurrence of lung cancer. Examination of the distribution of cases according to histological type showed a high proportion of adenocarcinomas (62%), which was consistent with the predominance of adenocarcinoma among non-smoking cases previously reported in the literature[10,11].

Analyses according to occupation and industry showed few significant results, but the elevated, although not significant, ORs observed for painters, warehouse porters, labourers and motor vehicle repairers are consistent with the literature[12]. On the other hand, some known associations were not found. In particular, we did not observe an increase in risk among welders or, more generally, among those who had performed welding activities, contrary to other studies[13]. Nor did we observe any association with exposure to asbestos. This result may be explained by misclassification and low exposure levels. Exposure was assessed independently of case-control status with an algorithm using questions on exposing tasks, but some tasks may have not been accurately reported. Exposure levels were probably low even if they were not formally assessed, as activities entailing high exposure levels such as the mining and milling of asbestos or the manufacture of asbestos-containing products were never present in the FWI.

The most striking finding of our study is the significantly increased risk of lung cancer in sugarcane farm workers and in sugarcane growing in general, as well as a non-significant increase in risk in rum manufacturing. Although the number of studies is limited, excess lung cancer risk has been observed in sugarcane growing or in the sugarcane industry[8,14,15]. This excess could be due to the presence of biogenic amorphous silica fibers in sugarcane leaves, which have characteristics similar to those of asbestos[16]. We found elevated ORs associated with cutting, crushing, collecting or handling the cane. These fibers can also be transformed into crystalline silica at high temperatures, during sugar or rum production or during the burning of cane before or after cutting. Another exposure that may arise from sugarcane burning is exposure to polycyclic aromatic hydrocarbons (PAHs), a known risk factor for lung cancer[17,18]. However, sugarcane burning was not associated with lung cancer risk in our study.

In contrast, the use of herbicide treatments in sugarcane was associated with an increased risk of lung cancer. Herbicide exposure in banana cultivation and to a lesser extent in other crops was also associated with an elevated risk of lung cancer. It was not possible to analyse chemical families or active substances, as the products used were not always clearly identified in the questionnaires, but rather by trade names, and not all exposed subjects could recall them. The most frequently cited herbicides are glyphosate, paraquat, phenoxy herbicides and triazines. Few studies have been conducted to assess the relationship between lung cancer and specific substances used as pesticides. The results come almost exclusively from the Agricultural Health Study[19] and do not show an association between lung cancer risk and the herbicides mentioned by the participants in our study. Conversely, associations between lung cancer risk and several insecticides used in the FWI have been suggested, including carbamates, organophosphates such as diazinon, and organochlorines such as dieldrin and lindane[19,20]. No clear association with insecticide exposure was however found in our study.

A limitation of our study is the relatively small number of cases and the consequently limited statistical power. Misclassification of exposure may also have occurred, which is likely to be non-differential. Self-reported occupational history is generally considered reliable, and coding was performed blind to case-control status. Exposures were assessed from self-reported information on tasks performed and materials handled, but the assessment was performed automatically regardless of case-control status. Data on the duration and frequency of exposure to certain tasks and substances were sometimes missing, and we were not able to assess exposure levels. This relatively crude exposure assessment combined with the small sample size precluded in-depth analyses by subgroups and the study of dose-response relationships.

Nevertheless, our study has several strengths. Selection bias is probably minimal. We were able to identify almost exhaustively the lung cancer cases that occurred within the study period. Participation rates were satisfactory for a population-based study and cases not included in the study were similar to included cases in terms of sex, age, and histological type. The control group was a random sample of the population with a distribution by socioeconomic status similar to that of the general population. We also used detailed questionnaires allowing us to identify a substantial amount of information on the subjects’ occupational history, aside from job titles alone. Finally, our study was able to examine occupational exposures in activities that can only be studied in a limited number of populations, such as sugarcane work or banana cultivation.

## CONCLUSION

Our study showed that occupational risk factors contributed to the occurrence of lung cancer in the FWI, and confirmed the role of specific exposures related to sugarcane work in lung cancer risk. Our findings highlighted the role of herbicides used in sugarcane cultivation and suggest more generally an association between exposure to herbicides and lung cancer.

## Data Availability

All data produced in the present study are available upon reasonable request to the authors

## Acknowledgements

We would like to thank our clinical research associates Lucina Lipau and Audrey Pomier for their help in data collection.

## Ethics approval

Written informed consent was provided by all participants in the study. Ethics approval was granted to this study by the French Data Protection Authority (CNIL, Commission Nationale de l’Informatique et des Libertés) no. DR-2015-2027; and the institutional review board of the French National Institute of Health and Medical Research (IRB INSERM n°01–036).

